# Prevalence and disease risks for male and female sex chromosome trisomies: a registry-based phenome-wide association study in 1.5 million participants of MVP, FinnGen, and UK Biobank

**DOI:** 10.1101/2025.01.31.25321488

**Authors:** Shanlee M. Davis, Aoxing Liu, Craig C. Teerlink, Dana M. Lapato, Bryan Gorman, Giulio Genovese, Madhurbain Singh, Mary P. Reeve, Amanda Elswick Gentry, Kati M. Donner, Timo P. Sipilä, Awaisa Ghazal, Meghana S. Pagadala, Matthew S. Panizzon, Eva E. Lancaster, FinnGen banner authorship, UKB working group authorship, Chris Chatzinakos, Andrea Ganna, Tim B. Bigdeli, Mark J Daly, Julie A. Lynch, Judith Ross, Roseann E. Peterson, Richard L. Hauger

**Affiliations:** Department of Pediatrics, School of Medicine, University of Colorado School of Medicine, Aurora, CO, USA; eXtraOrdinarY Kids Clinic, Children’s Hospital Colorado, Aurora, CO, USA; Analytic and Translational Genetics Unit, Massachusetts General Hospital, Boston, MA, USA; Program in Medical and Population Genetics, Broad Institute of Harvard and MIT, Cambridge, Massachusetts, USA; Stanley Center for Psychiatric Research, Broad Institute of MIT and Harvard, Cambridge, MA, USA; Center for Genomic Medicine, Massachusetts General Hospital, Boston, MA, USA; Institute for Molecular Medicine Finland (FIMM), University of Helsinki, Helsinki, Finland; VA Informatics and Computing Infrastructure (VINCI), VA Salt Lake City Health Care System, Salt Lake City, UT, USA; Division of Epidemiology, Department of Internal Medicine, University of Utah School of Medicine, Salt Lake City, UT, USA; Department of Human & Molecular Genetics, Virginia Institute for Psychiatric and Behavioral Genetics, Virginia Commonwealth University, Richmond, Virginia, USA; VA Boston Healthcare System, Boston, MA, USA; Department of Psychiatry, Virginia Institute for Psychiatric and Behavioral Genetics, Virginia Commonwealth University, Richmond, Virginia, USA; Research Service, VA San Diego Healthcare System, San Diego, CA, USA; Medical Scientist Training Program, University of California San Diego, La Jolla, CA, USA; Biomedical Science Program, University of California San Diego, La Jolla, CA, USA; Department of Psychiatry and Behavioral Sciences, Institute for Genomics in Health, SUNY Downstate Health Sciences University, Brooklyn, NY; VA New York Harbor Healthcare System, Brooklyn, NY; Department of Epidemiology and Biostatistics, School of Public Health, SUNY Downstate Health Sciences University, Brooklyn, NY; Nemours Children’s Hospital DE, Wilmington, DE, USA; Department of Pediatrics, School of Medicine, Thomas Jefferson University, Philadelphia, PA, USA; Center of Excellence for Stress and Mental Health (CESAMH), VA San Diego Healthcare System, San Diego, CA, USA; Center for Behavior Genetics of Aging, School of Medicine, University of California San Diego, La Jolla, CA, USA

## Abstract

Sex chromosome trisomies (SCT) are the most common whole chromosome aneuploidy in humans. Yet, our understanding of the prevalence and associated health outcomes is largely driven by observational studies of clinically diagnosed cases, resulting in a disproportionate focus on 47,XXY and associated hypogonadism. We analyzed microarray intensity data of sex chromosomes for 1.5 million individuals enrolled in three large cohorts—Million Veteran Program, FinnGen, and UK Biobank—to identify individuals with 47,XXY, 47,XYY, and 47,XXX. We examined disease conditions associated with SCTs by performing phenome-wide association studies (PheWAS) using electronic health records (EHR) data for each cohort, followed by meta-analysis across cohorts. Association results are presented for each SCT and also stratified by presence or absence of a documented clinical diagnosis for 47,XXY. We identified 2,769 individuals with (47,XXY: 1,319; 47,XYY: 1,108; 47,XXX: 342), most of whom had no documented clinical diagnosis (47,XXY: 73.8%; 47,XYY: 98.6%; 47,XXX: 93.6%). The identified phenotypic associations with SCT spanned all PheWAS disease categories except neoplasms. Many associations are shared among three SCT subtypes, particularly for vascular diseases (e.g., chronic venous insufficiency (OR [95% CI] for 47,XXY 4.7 [3.9,5.8]; 47,XYY 5.6 [4.5,7.0]; 4 7,XXX 4.6 [2.7,7.6], venous thromboembolism (47,XXY 4.6 [3.7-5.6]; 47,XYY 4.1 [3.3-5.0]; 47,XXX 8.1 [4.2-15.4]), and glaucoma (47,XXY 2.5 [2.1-2.9]; 47,XYY 2.4 [2.0-2.8]; 47,XXX 2.3 [1.4-3.5]). A third sex chromosome confers an increased risk for systemic comorbidities, even if the SCT is not documented. SCT phenotypes largely overlap, suggesting one or more X/Y homolog genes may underlie pathophysiology and comorbidities across SCTs.

## Introduction

Sex chromosome trisomies (SCT), characterized by the presence of an additional copy of chromosome X (47,XXY and 47,XXX) or chromosome Y (47,XYY), are the most common type of chromosomal aneuploidies in humans, with an estimated prevalence ∼0.2% live births worldwide.^1–4^ Most studies thus far have focused on clinically diagnosed cases and, therefore, have relatively small sample sizes and have primarily investigated 47,XXY (Klinefelter syndrome). Males with 47,XXY have been typically described as having tall adult stature, high body fat percentage, microorchidism, gynecomastia, azoospermia, and learning disabilities.^5,6^ Clinical studies have shown that Klinefelter syndrome is often comorbid with infertility, diabetes, osteoporosis, metabolic syndrome, and psychiatric disorders, among many other chronic conditions.^5,7^

Although Klinefelter syndrome is the most diagnosed SCT, approximately three-quarters of those with 47,XXY are undiagnosed.^2–4^ The failure to include undiagnosed individuals with SCT, particularly those with the rarely diagnosed SCT subtypes 47,XYY and 47,XXX, in research cohorts introduces multiple biases, resulting in only those with certain clinical features being formally diagnosed and studied. The rich genomic data available in large biobanks allow systematic identification of SCTs genetically and, thus, provide a unique opportunity for a comprehensive exploration of SCTs, irrespective of clinical diagnosis status. For example, the analysis of genotyping data of 200,000 men of European ancestry in the UK Biobank confirms that male SCTs are mostly unrecognized and have high risks for psychiatric disorders and cardiovascular disorders; additionally, the study suggests that reproductive dysfunction is more specific to 47,XXY than to 47,XYY.^2^ However, such findings are limited to male SCTs, with conditions for females remaining underexplored. Recently, a multicohort study of 642,000 adults combining data of UK Biobank and the US Geisinger MyCode Community Initiative reported an increased risk of venous thromboembolism (VTE) for all three SCT subtypes.^3^ A study on the iPSYCH2015 case-cohort dataset (N=119,000) showed that each SCT is associated with at least one index psychiatric disorder with no significant difference seen between clinically diagnosed and undiagnosed SCTs.^8^ Although some comorbidities are seen across different SCT subtypes, most research has studied SCTs individually; studies assessing shared phenotypes across SCTs have generally focused on developmental/psychological domains as opposed to physical health domains and have typically been focused on the ∼15% of individuals with SCTs that have a clinical diagnosis. Additionally, these previous studies primarily include individuals of European ancestry; no study has assessed how generalizable these findings are across populations, the phenome, or different clinical practice systems.

Here, through international collaborative efforts, we aim for a comprehensive understanding of the prevalence of disease risks for individuals with genotype-identified SCTs in both genetic males and females by assessing the heterogeneity of disease associations across SCT subtypes, SCT clinical diagnosis status, and nations. We identified individuals with SCTs in three large cohorts including a total of 1.5 million individuals, and then performed phenome-wide association studies (PheWAS) to assess disease associations for each SCT individual compared to matched controls. This study serves as a foundation for more in-depth analyses of SCT phenotypes in population-based biobanks.

## Methods

### Study participants

This retrospective cohort study considered 1.5 million individuals enrolled in three large cohorts—the Million Veteran Program (MVP), FinnGen, and UK Biobank—all of which have participants genotyped using blood-derived DNA and linked to detailed electronic health records (EHR). The Veterans Affairs (VA) MVP, a US-based biobank started in 2011, is one of the largest genetic biobanks and is linked to rich, longitudinal EHR in the VA healthcare system. The MVP cohort used in this study included approximately ∼650,000 Veterans; recruitment, follow-up, and primary data collection is ongoing.^9^ FinnGen is a public-private partnership research project in Finland that is investigating the genome and digital EHR data of 500,000 Finns. FinnGen phenotype data derive from diverse national health registers including diagnoses from hospitalization, healthcare reimbursement, and causes of death which began at or before 1969; the notable fraction of hospital-based recruitment has led to enriched disease endpoints in FinnGen.^10^ The UK Biobank is a prospective cohort in the United Kingdom, started in 2006, with de-identified genomic, medical, and self-reported phenotypic data on approximately 500,000 volunteer participants aged 40 to 70.^11^ Eligible participants were recruited by invitation from population-based registries between 2006-2010, with continued longitudinal follow-up ongoing.

### Participant consent

All MVP participants provided written informed consent, and the study was approved by the VA Central Institutional Review Board (IRB). FinnGen was approved by the Coordinating Ethics Committee of the Hospital District of Helsinki and Uusimaa (HUS), and patients and control subjects provided informed consent for biobank research, based on the Finnish Biobank Act. All UK Biobank participants provided informed consent in accordance with the UK Biobank Ethics and Governance Framework (EGF), as approved by the UK Biobank Ethics and Governance Council (EGC), established by the UK Biobank funders, the Wellcome Trust, and the Medical Research Council.

### Exposures

The identification of SCT status was conducted within each participating cohort. For each cohort, we analyzed raw microarray intensity data from chromosome X and chromosome Y probes. We considered individuals with detectable Y chromosome intensity signals to be genetic males and those without to be genetic females. Then, we clustered participants into the following groups: 46,XY, 47,XXY, and 47,XYY (genetic males), 46,XX and 46,XXX (genetic females), or ambiguous, following the thresholds used by previous studies.^2,4,12^

To quantify the diagnosis rates for each SCT, we identified individuals with a clinical diagnosis if, at any time, their EHR included a code from the International Classification of Diseases, Ninth Revision (ICD-9) beginning with 758 or the International Classification of Diseases, Tenth Revision (ICD-10) Q97-99.^3,4^

### Outcomes

We used phecodes to represent an individual’s medical conditions for specific diseases. Within each cohort, we mapped ICD-9 and ICD-10 codes to 1,866 unique phecodes using the PheWAS catalog map v1.2.^13,14^ The phecodes were collapsed into one of 18 system-based categories akin to ICD chapters (i.e., cardiovascular disease, cancer, neurological disease, mental health and psychiatric disorders, gastrointestinal disease, kidney disease, liver disease, endocrine and metabolic disorders, respiratory disease, musculoskeletal disease, infections and parasitic diseases, pregnancy and reproductive disorders, skin and subcutaneous tissue disorders, blood and immune disorders, hematological and coagulation disorders, genitourinary disorders, eye disease, other conditions).

### Statistical analyses

To test the association between a given SCT subtype and lifetime risk of a disease, identified by phecodes, we adopted a matched case-control design. Each individual with a genetically-identified SCT was matched to five controls based on genetic sex, birth year, and genetic ancestry (when applicable). Logistic regression using the base R package was performed for each SCT case-control matched set using the identified SCT status as the exposure, a specific phecode as the binary outcome, and birth year, genetic ancestry (Admixed American, African, East Asian, European, unavailable), and the top ten genetic principal components as covariates. For each phecode outcome, the beta estimate, standard error, P value, and odds ratio (OR) with 95% confidence intervals were computed. For all analyses conducted within a cohort, a minimum of five individuals was required in each cell of the 2×2 contingency table (SCT present/absent, phecode present/absent) for any analyzed SCT subtype. The 47,XXY cohort had a sufficient number of clinically diagnosed cases, allowing for sensitivity analyses based on clinical diagnosis status.

The meta-analysis across three cohorts was performed with a fixed-effect model applied in the R meta package.^15^ The significance threshold was determined using the Bonferroni correction. The heterogeneity across cohorts was assessed with a Cochran’s Q test. Volcano plots were generated to visually display the PheWAS results from the three-cohort meta-analysis.

## Results

### Study participants

Among the 1.5 million individuals assessed, we identified 2,769 individuals (0.19%) with any SCT (186 per 100,000)—1,319 as 47,XXY, 1,108 as 47,XYY, and 342 as 47,XXX. The majority (86.2%) were undiagnosed and presumably unaware of their SCT; the clinical diagnosis rates were 26.2% for 47,XXY, 1.4% for 47,XYY, and 6.4% for 47,XXX (see Table 1 for demographic details by cohort).

**Table 1.** Demographics of individuals with sex chromosome trisomies (SCTs) in three population-based datasets.

| Database | MVP | UK Biobank | FinnGen (DF10) | All 3 Datasets |
| --- | --- | --- | --- | --- |
| Total Population | n=654,619 | n=488,377 | n=342,193 | N=1,485,189 |
| Sex, % male | 91% | 46% | 43% | 65% |
| Sex chromosome trisomies (n) | 1,670 | 514 | 584 | 2,769 |
| 47,XXY | 862 | 225 | 232 | 1,319 |
| 47,XYY | 747 | 166 | 195 | 1,108 |
| 47,XXX | 61 | 123 | 157 | 342 |
| Prevalence of all SCTs (per 100,000) | 255 | 105 | 171 | 186 |
| 47,XXY prevalence (per 100,000 males) | 145 | 100 | 158 | 136 |
| 47,XYY prevalence (per 100,000 males) | 125 | 74 | 133 | 115 |
| 47,XXX prevalence (per 100,000 females) | 104 | 47 | 80 | 66 |
| Genetic Ancestry |  |  |  |  |
| European | 1,399 (83.8%) | 487 (94.6%) | 584 (100%) | 2470 (89.2%) |
| African | 141 (8.4%) | 9 (1.8%) | 0 | 150 (5.4%) |
| Admixed American | 85 (5.1%) | 0.8% | 0 | 85 (3.1%) |
| East Asian | 25 (1.5%) | 2.8% | 0 | 25 (0.9%) |
| Unavailable | 19 (1.1%) | 0 | 0 | 19 (0.7%) |
| Age at last follow-up (yrs)* | 68.0 (14.2) | 55.3 (8.3) | 56.6 (18.8) | -- |
| Clinical SCT diagnosis | 245 (14.7%) | 49 (9.5%) | 88 (15.1%) | 382 (13.8%) |
| 47,XXY | 226 (26.2%) | 49 (21.8%) | 70 (30.2%) | 345 (26.2%) |
| 47,XYY | 2 (0.1%) | 0 | 13 (6.7%) | 15 (1.4%) |
| 47,XXX | 17 (27.9%) | 0 | 5 (3%) | 22 (6.4%) |
| Data are shown as mean ± standard deviation (SD) for numerical data, and n (%) for categorical data, unless otherwise specified. MVP = Million Veteran Program; UK = United Kingdom; EHR = electronic health record. | | | | |
| *Age of recruitment in UK Biobank |  |  |  |  |

### SCT PheWAS analysis

Of the 1,866 phecodes used for ICD code mapping, 1,182 unique phecodes were available for use in our PheWAS meta-analysis, with 452 for all SCTs and 516 for two of the three SCTs. After Bonferroni correction, we identified 463 SCT-phecode associations for 283 unique phecodes. The identified associations covered all examined disease categories, except for neoplasms (Figures 1-4, Tables S1-S3).

**Figure 1.**
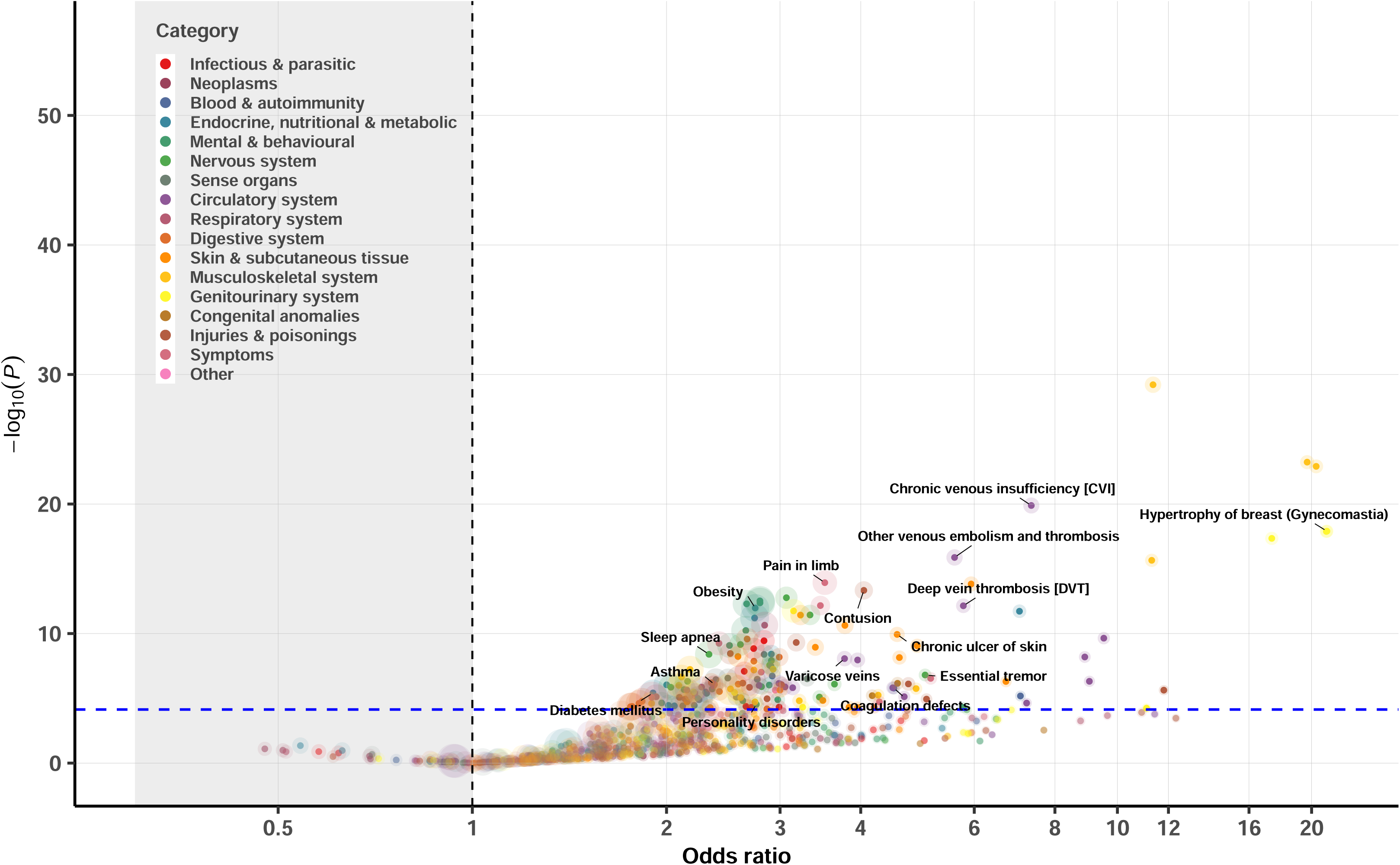
Association analysis of disease diagnosis for 1,319 genetically identified 47,XXY from a meta-analysis of MVP, FinnGen, and UK Biobank. Each 47,XXY case was matched to five 46,XY controls based on birth year and genetic ancestry (when applicable). Logistic regression was used to examine the association between the 47,XXY status and a given phecode within each cohort, followed by a fixed-effect meta-analysis across cohorts. Dashed lines denote the statistically significant threshold after Bonferroni correction (P=0.05/1,076=4.6e-05); colors represent disease categories; the size of the circle reflects the N of disease cases analyzed for each phecode.

**Figure 2.**
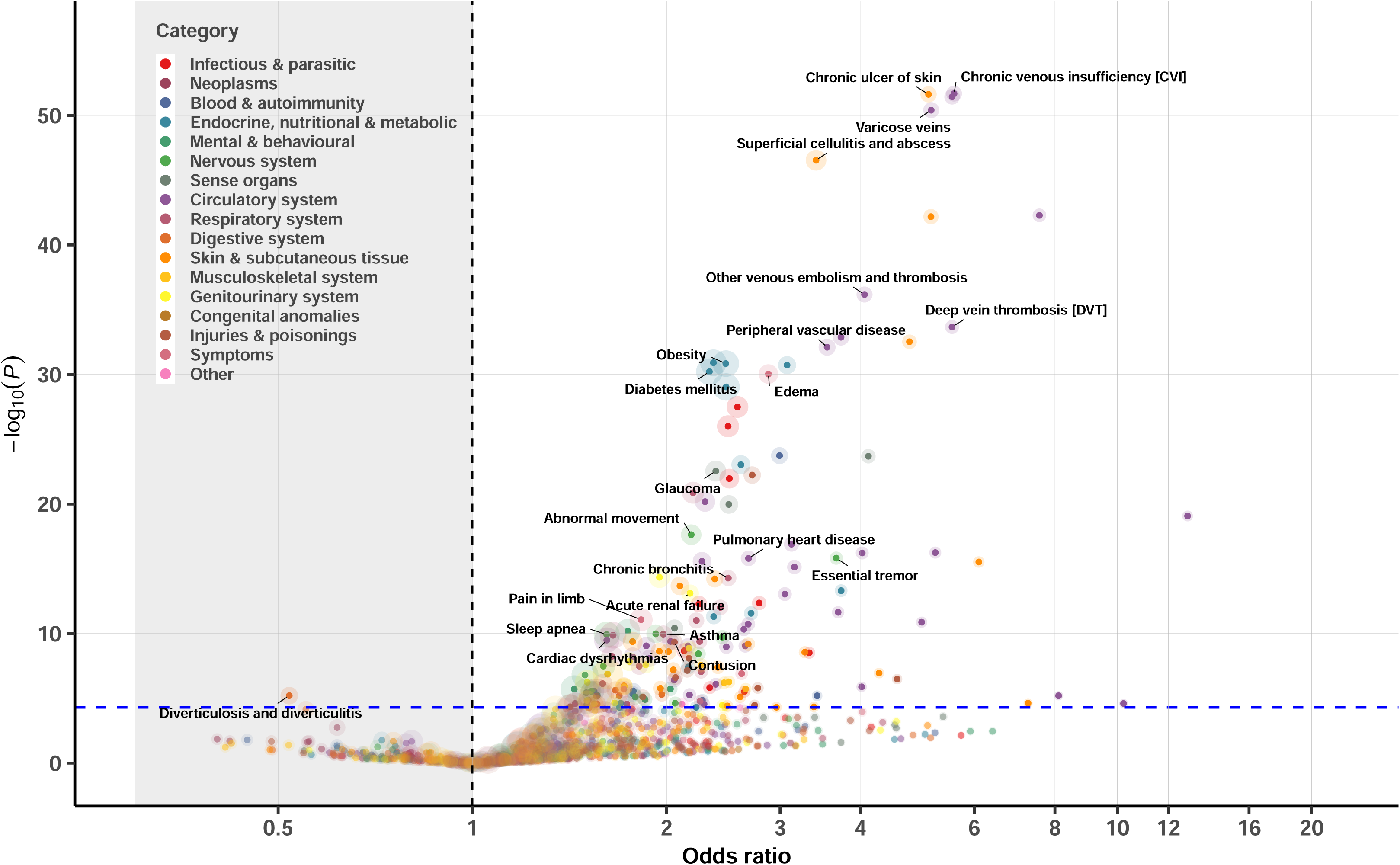
Association analysis of disease diagnosis for 1,108 genetically identified 47,XYY from a meta-analysis of MVP, FinnGen, and UK Biobank. Each 47,XYY case was matched to five 46,XY controls based on birth year and genetic ancestry (when applicable). Logistic regression was used to examine the association between the 47,XYY status and a given phecode within each cohort, followed by a fixed-effect meta-analysis across cohorts. Dashed lines denote the statistically significant threshold after Bonferroni correction (P=0.05/1,007=5.0e-05); colors represent disease categories; the size of the circle reflects the N of disease cases analyzed for each phecode.

**Figure 3.**
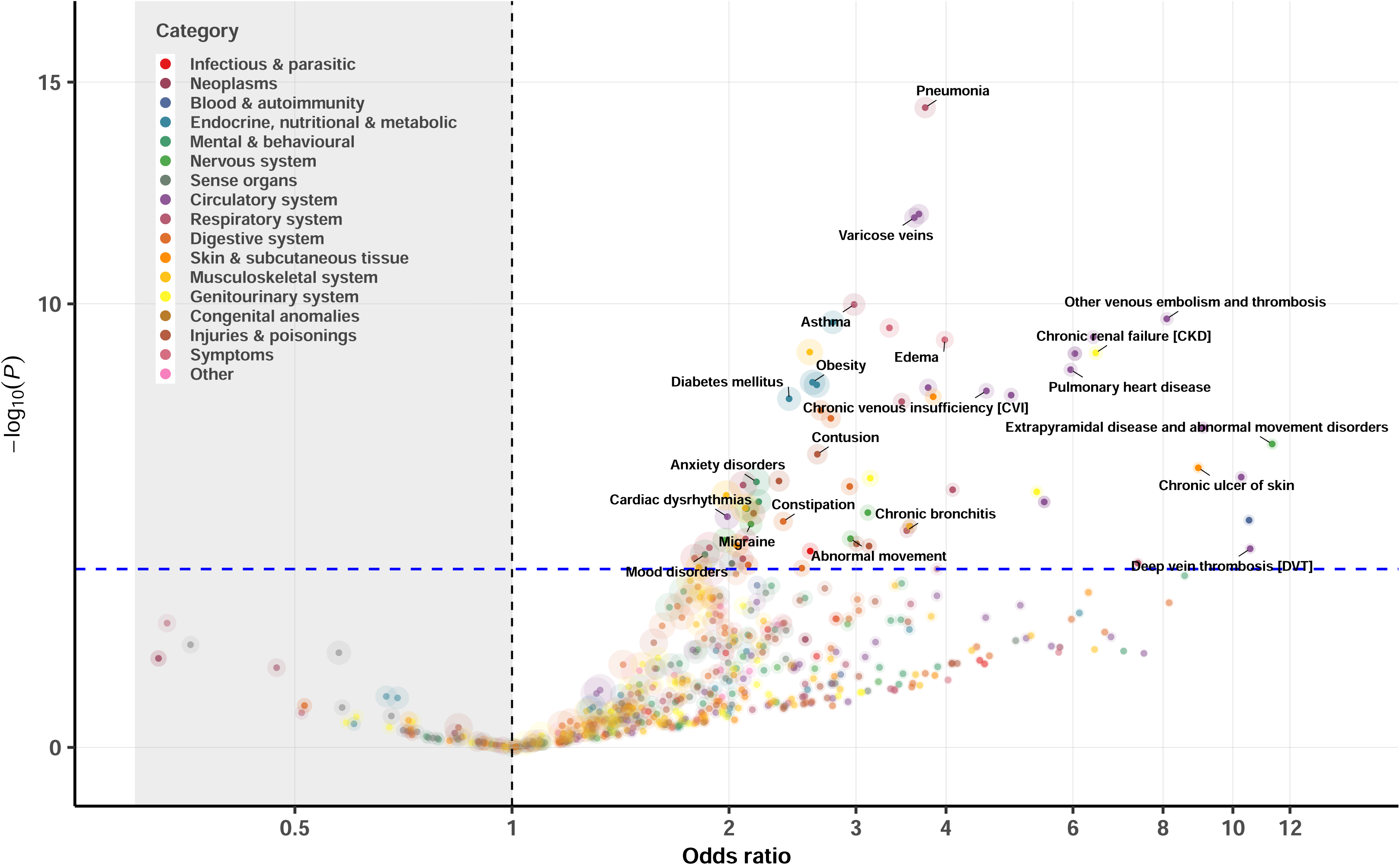
Association analysis of disease diagnosis (represented by phecodes) for 342 genetically identified 47,XXX from a meta-analysis of MVP, FinnGen, and UK Biobank. Each 47,XXX case was matched to five 46,XX controls based on birth year and genetic ancestry (when applicable). Logistic regression was used to examine the association between the 47,XXX status and a given phecode within each cohort, followed by a fixed-effect meta-analysis across cohorts. Dashed lines denote the statistically significant threshold after Bonferroni correction (P=0.05/519=9.6e-05); colors represent disease categories; the size of the circle reflects the N of disease cases analyzed for each phecode.

**Figure 4.**
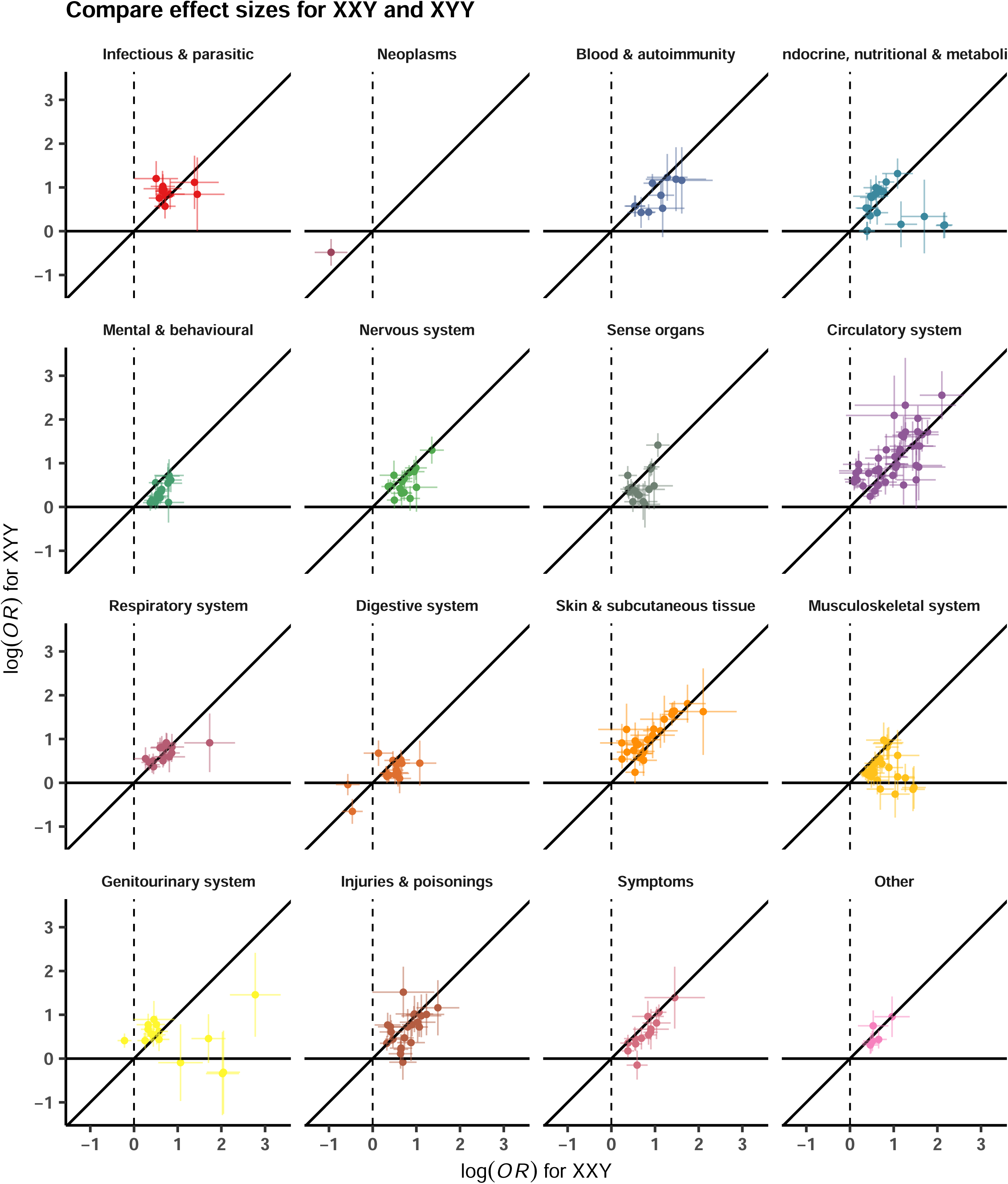
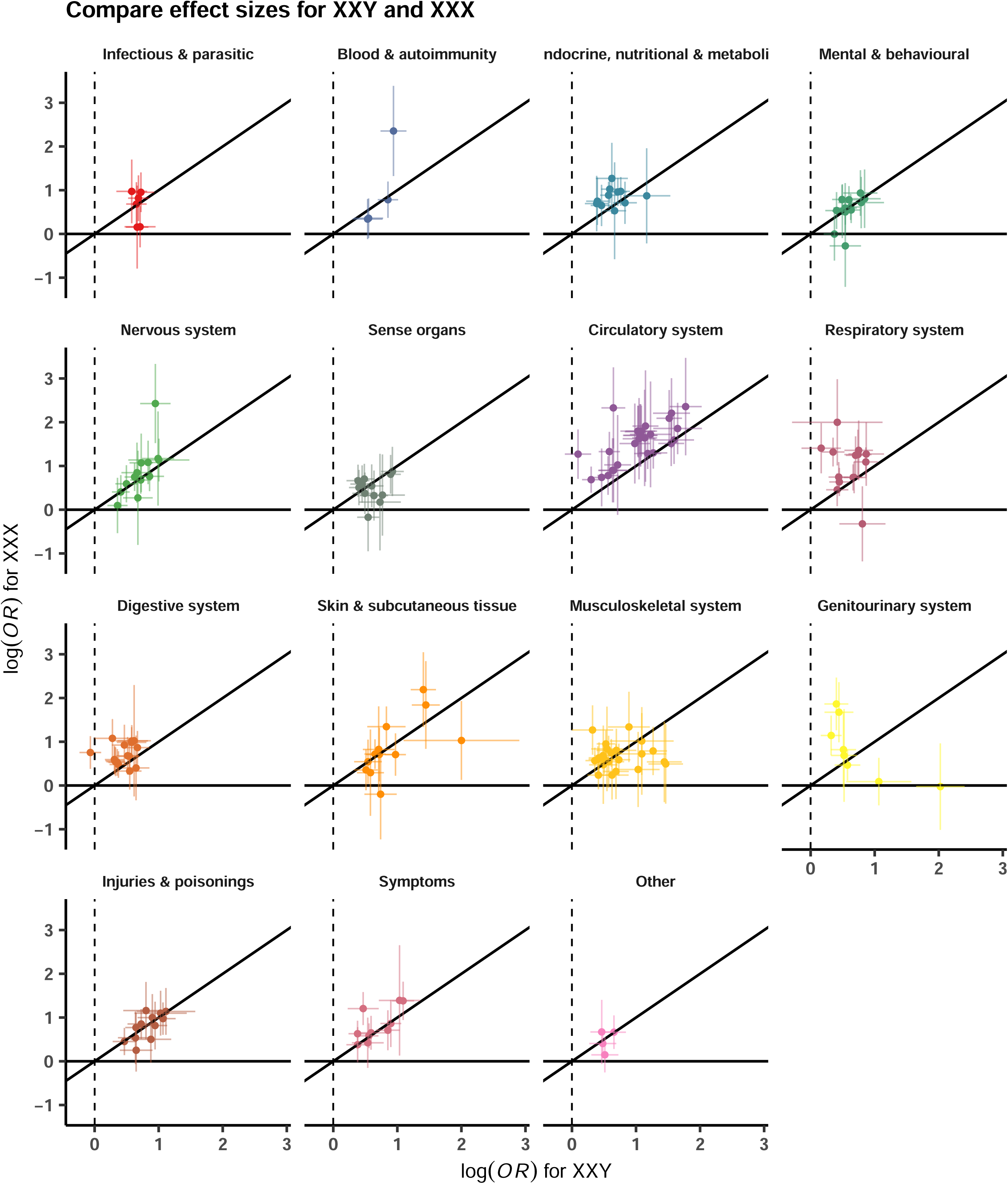
Association of disease diagnosis across SCT subtypes. Panel (a) depicts the comparison of disease associations of two male SCT subtypes - 47,XXY (n=1,319) and 47,XYY (n=1,108), and panel (b) for two SCT subtypes with an extra X chromosome among genetic males (47, XXY) (n=1,319) and genetic females (47,XXX) (n=342). The odds ratios are from a meta-analysis of MVP, FinnGen, and UKB. Only disease diagnoses exerting significant association (after Bonferroni correction) for at least one examined SCT subtype are considered. Colors represent disease categories.

Notably, 43 phecodes were significantly associated with all three SCTs. Among these, 16 were diseases of the circulatory system, 7 were diseases of the respiratory system, and 4 were metabolic disorders (Table 2). The associations with circulatory system-related diseases were particularly strong for vascular diseases including the following: (a) cerebrovascular disease (ORs from 1.4 [1.2-1.7] for 47,XYY to 2.2 [1.4-3.4] for 47,XXX, P value for heterogeneity=0.12), (b) atherosclerosis (ORs from 2.1 [1.6-2.7] for 47,XYY to 4.5 [1.8-11.3] for 47,XXX, P value for heterogeneity=0.15), (c) other venous embolism and thrombosis (ORs [95% CI] from 4.1 [3.3-5.0] 47,XYY to 8.1 [4.2-15.4] for 47,XXX, P value for heterogeneity=0.13), (d) chronic venous insufficiency (ORs from 4.6 [2.7-7.6] for 47,XXX to 5.6 [4.5-7.0] for 47,XYY, P value for heterogeneity=0.50), and (e) deep vein thrombosis (ORs from 5.5 [4.2-7.3] for 47,XYY to 10.6 [3.5-32.2] for 47,XXX, P value for heterogeneity=0.54). Examples of other conditions that were significantly more prominent in all three SCT groups include obesity, type 2 diabetes, dermatophytosis, atopic dermatitis, asthma and chronic airway obstruction, sleep apnea, anemias, abnormal movement, peripheral nerve disorders, essential tremor, glaucoma, edema, chronic skin ulcers, syncope, dysphagia, and cholelithiasis.

**Table 2.**
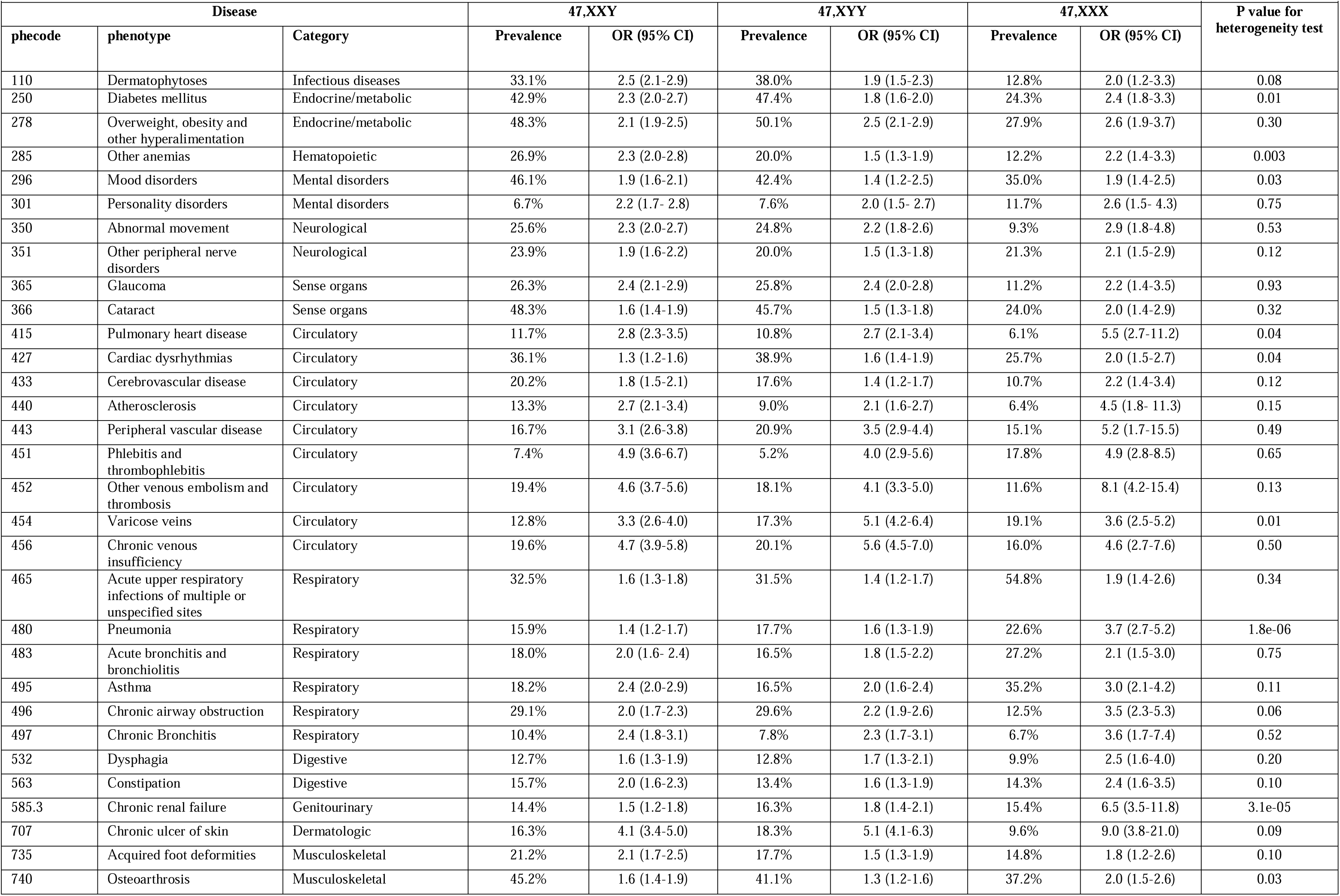
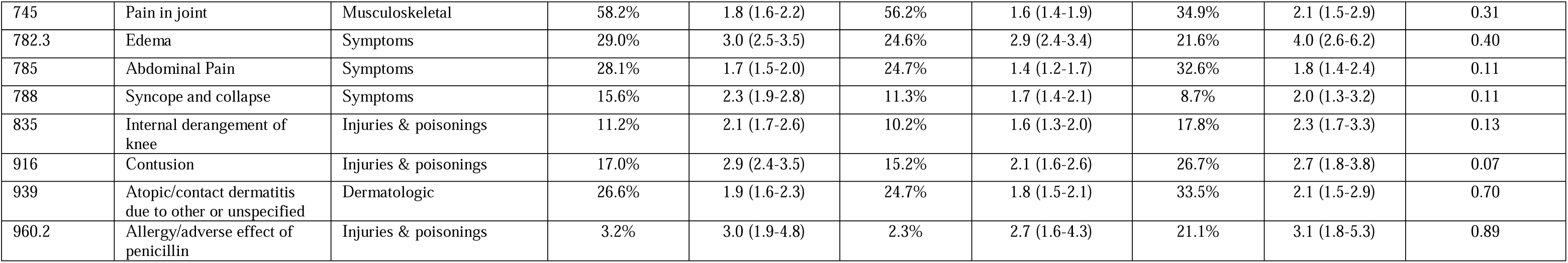
Select phenotypes with universally higher prevalence among all SCT conditions compared to matched controls.

Anxiety disorders were twice as common in individuals with an additional X chromosome compared to controls (47,XXY: 1.8 [1.6,2.1]; 47,XXX: 2.2 [1.6,3.0]), but this association was not nearly as strong in those with 47,XYY (1.2 [1.1, 1.4]). Males with SCT, but not females, had a higher odds of diseases of the sebaceous glands (47,XXY: 1.7 [1.4,2.0]; 47,XYY: 2.1 [1.7,2.5]), cellulitis (47,XXY: 2.6 [2.2,3.1]; 47,XYY: 3.4 [2.9,4.1]), and urinary tract infections (47,XXY: 1.8 [1.5,2.2]; 47,XYY: 1.9 [1.5,2.3]). Conditions with higher odds unique to 47,XXY include testicular dysfunction (8.6 [7.1,10.2]), infertility (16.1 [9.0,28.7]), gynecomastia (7.7 [5.3,11.3]), and osteoporosis (4.3 [3.3,5.7]). Females with 47,XXX had a disproportionately higher odds of chronic renal failure (6.5 [3.5,11.8]) and pneumonia (3.7 [2.7,5.2]). No unique phenotype risk was identified in 47,XYY.

### Stratification by diagnosis status in EHR

Among those with the 47,XXY SCT in the cohort, men with a clinical diagnosis of 47,XXY/Klinefelter syndrome in their EHR (345/1,319; 26.2%) were more likely than those without an EHR diagnosis to have diagnoses mapping to phecodes for testicular dysfunction, anterior pituitary dysfunction (likely miscoded as hypogonadotropic hypogonadism rather than hypergonadotropic hypogonadism), infertility, osteoporosis, gynecomastia, and sleep disorders (Figure 5, Figure S1, Figure S2, Table S4). Similar analyses among the 47,XYY and 47,XXX karyotype groups could not be performed because too few individuals had a clinical diagnosis in their EHR (Table 1).

**Figure 5.**
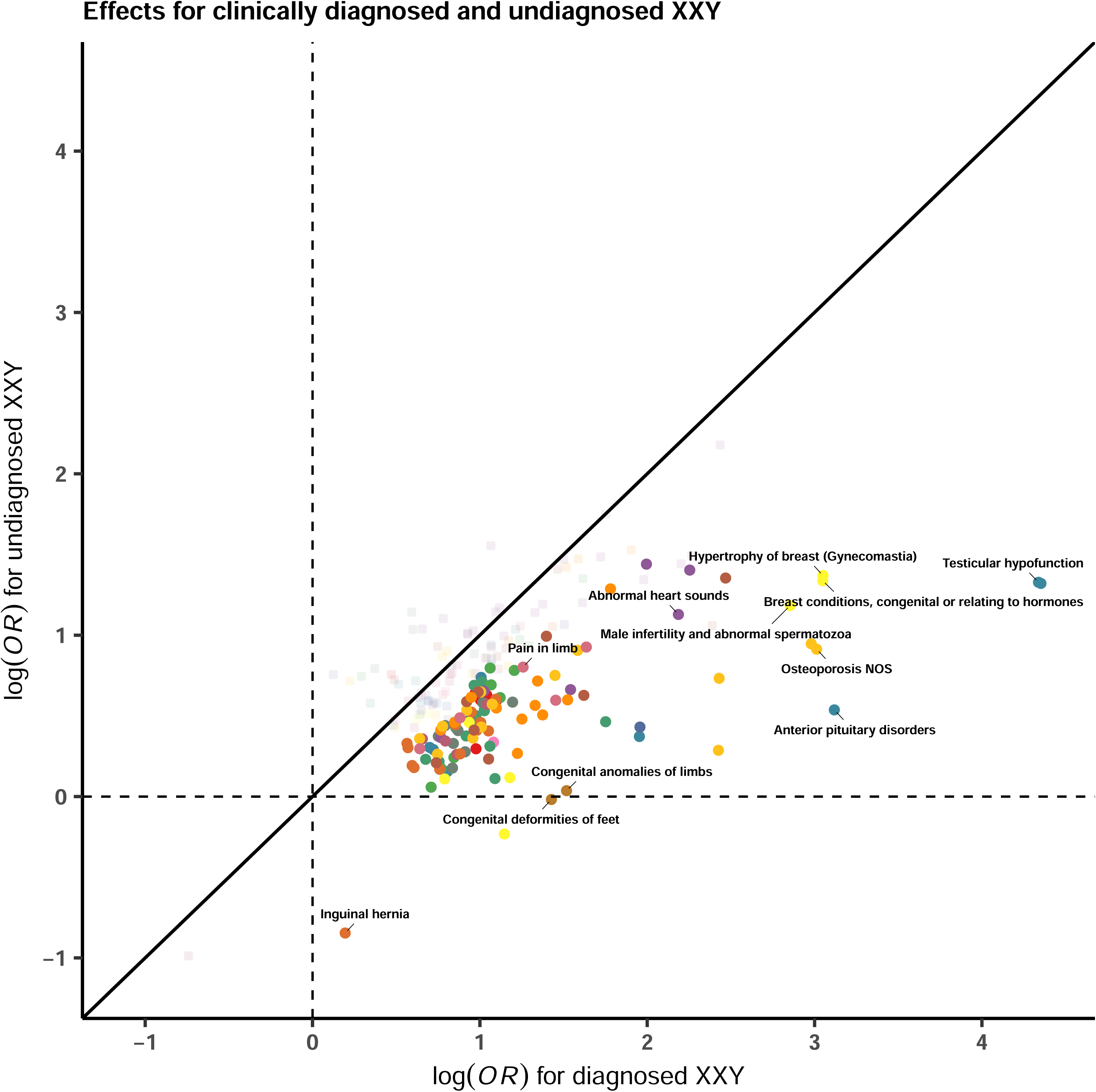
Odds ratios of disease diagnosis for 47,XXY from clinically diagnosed cases (n=345) versus clinically undiagnosed cases (n=974). The odds ratios are from a meta-analysis of MVP, FinnGen, and UKB. Only disease diagnoses exerting significant association (after Bonferroni correction) for at least one examined SCT subtype are considered. Colors represent disease categories. Only disease diagnoses that exert different effects between two examined SCT subtypes at a nominal P value from a two-sided t-test are colored.

## Discussion

This international, collaborative study of nearly 1.5 million participants within three distinct population-based genomic biorepositories yields the largest genotype-phenotype analysis of individuals with SCT conditions. Our results confirm ascertainment estimates from previous studies, showing that ∼75% of males harboring an additional X chromosome and >90% of individuals with 47,XYY and 47,XXX are not clinically identified, even late into adulthood. Compared to matched controls, individuals with SCT had a higher prevalence of numerous chronic cardiovascular and neuropsychiatric conditions that have historically been associated with SCT. In addition, the large sample size and comprehensive PheWAS approach in our study allowed for both greater precision and identification of novel observations that extend the existing literature. This work emphasizes that the systemic phenotype of an additional sex chromosome is remarkably similar across genetic sexes, ancestries, and nations. We propose that increased pseudoautosomal and/or X/Y homolog gene dosage underlies much of the observed SCT phenotype, rather than hypogonadism isolated to XXY.^16–19^

The majority of the SCT literature is limited to 47,XXY/Klinefelter syndrome, reflecting the large discrepancy in clinical ascertainment, likely due to the high prevalence of testicular dysfunction in 47,XXY and the lack of pathognomonic features in 47,XYY or 47,XXX. Historically, the comorbidities of the 47,XXY phenotype have been attributed to testosterone deficiency, with appropriate testosterone hormone replacement promising to mitigate these sequelae. The findings of our study, however, add to the growing body of research minimizing the causative role of testosterone deficiency on the systemic phenotypes of 47,XXY, and instead suggesting that most of the phenotype is secondary to the presence of an additional X chromosome. Notable exceptions to this include infertility, gynecomastia, and osteoporosis which were associated with 47,XXY (both diagnosed and undiagnosed) but not 47,XYY. Aside from these few exceptions, our results suggest that all three SCT conditions could be pooled together for many clinical outcomes, increasing both feasibility and generalizability for future investigations of the underlying pathophysiology and targeted interventions across SCTs.^20,21^

The concerning vascular phenotype we observed in all three SCTs is consistent with a recent study of VTE within the UK Biobank and MyCode datasets, which found a higher prevalence and 10-year incidence of VTE events among 733 persons with SCT compared to euploid males and females.^3^ Odds ratios for VTE history in SCT groups for that study ranged from 4.0-7.8, essentially the same as the pooled estimate that we found (ORs 4.1-8.1). While the 47,XXY karyotype has been associated with a hypercoagulable state for many decades, the VTE risk for individuals with 47,XYY and 47,XXX karyotypes has only recently been suggested. In addition to VTE, the higher odds of peripheral vascular disease, chronic venous insufficiency, varicose veins, and skin ulcerations indicate that vascular disease pathophysiology in SCTs is not isolated to coagulopathy. One potential contributor to the observed vascular pathophysiology is the high prevalence of diabetes and insulin resistance across the three SCTs, which may involve amplification of gene dose in the pseudoautosomal region (PAR) of both the X and Y chromosome.^22,23^ Intriguingly, several other conditions found to be more common in persons with SCTs could also have an underlying vascular pathology, including glaucoma, neuropsychiatric and neurodegenerative disorders, vasovagal syncope, and even primary gonadal dysfunction.^24–26^ These results emphasize the need to further investigate the unique, hormone-independent, vascular pathophysiology accompanying SCT. Noted in other studies and reinforced by our data, SCT does not confer an increased risk of hypertension or ischemic heart disease.^27,28^

Several findings of this study were unexpected based on prior literature. First, across the three cohorts, glaucoma was twice as common in individuals with SCT compared to age-matched controls, affecting 24% of all individuals with SCT. Ophthalmologic abnormalities have been described in 47,XXY.^29–31^ However, glaucoma has not been specifically reported, and certainly not at the high prevalence found within these cohorts. Therefore, glaucoma may be an important age-related SCT comorbidity for which surveillance in individuals with SCT is warranted. Next, although it has been reported that autoimmunity is higher in SCTs with an additional X chromosome,^32–34^ our analysis failed to confirm this observation. The point estimate for the prevalence of the most common autoimmune condition, hypothyroidism, was actually lower for all three SCTs compared to previous studies. Although this was of borderline statistical significance, observation across all three datasets adds confidence that there is no increased risk for hypothyroidism, contrary to previous studies.^35^ Other autoimmune conditions such as celiac disease, psoriasis, rheumatoid arthritis, inflammatory bowel disease, multiple sclerosis, and lupus were rare and were not more prevalent in MVP, FinnGen, and UKBiobank participants with SCT. Similarly, while breast cancer has been reported to be a significant risk in men with 47,XXY,^36^ it was not possible to assess this association given there were fewer than five breast cancer cases in all three datasets, representing <3% of persons with 47,XXY. These data suggest that prior research on clinically ascertained individuals with SCT may overestimate risk for certain conditions, which has implications for the screening recommendations for those with SCT. Finally, whether SCT may confer a protective effect against certain conditions, particularly neoplasms, warrants further investigation.

These results highlight the importance of developing and testing targeted interventions to prevent individuals with SCT from acquiring systemic comorbidities with aging. To date, intervention studies in SCT conditions almost exclusively evaluate testosterone replacement therapy in hypogonadal men with 47,XXY. Notably, only a minority of those with confirmed SCT in these cohorts were clinically identified, and the mean age of diagnosis for the minority who were diagnosed was 60 years of age. Early diagnosis of SCT, which is now widely available with genetic testing methods, presents an opportunity to reduce morbidity from preventable conditions. For example, the lifetime prevalence of 20% for VTE conferred by SCTs could potentially be ameliorated by lifestyle (e.g., exercise, compression) and pharmacologic (e.g., aspirin, anticoagulants) interventions. As a final note, despite the differences highlighted within, the diagnoses affecting the highest proportion of individuals with SCT are akin to the general aging population; presbyopia and cataracts, periodontal diseases, hearing loss, hypertension, obesity, joint pain, and depression affect at least a third of aging individuals with SCTs, similar to their peers. These common conditions should be taken into clinical consideration and must not be disregarded when caring for individuals with SCT.

This study has several limitations. Our diverse, multinational PheWAS identified associations between SCTs and several clinical phenotypes but did not test for associations that may mediate these relationships, such as lifestyle and environmental exposures. Future analyses should validate specific phenotypic findings while controlling for applicable covariates and/or progression over time. Next, phecodes only represent conditions documented in the EHR, which will not detect phenotypic differences in SCTs that do not present as medical pathology. Neither subclinical comorbidities nor potentially advantageous traits associated with an additional X or Y chromosome could be identified. In addition, our approach compares the presence or absence of a particular phenotype and does not account for the severity of a particular phenotype. Additional analyses that account for age at presentation and/or number of times a phenotype is documented in the EHR may provide ways of assessing for phenotypic severity within an EHR dataset, but still may not encompass the severity of the condition or the impact on quality of life. Future research could also integrate multi-omic data (e.g., epigenomics, metabolomics, and proteomics) to uncover underlying biological pathways that influence the phenotypic outcomes of SCT. Incorporating advanced machine learning models could help predict the progression of certain phenotypes over time and account for phenotypic severity more comprehensively.

Another potential limitation is that each of the three biobanks has ascertainment biases: MVP involves military service with its unique exposures; UK Biobank has a previously noted healthy, highly educated volunteer bias; FinnGen is enriched for disease endpoints and reflects a founding bottleneck.^10,37,38^ Nevertheless, our findings are not dissimilar from the large body of the existing literature, which has a bias toward clinically diagnosed individuals. Weighting statistical models to adjust for known biases could improve the robustness of results.

Moreover, employing meta-analytic techniques that account for heterogeneity between cohorts may help balance biases across different data sources. Finally, our analysis does not account for differences in ICD code use across the three healthcare systems or for loss of information when mapping ICD codes to phecodes.

Finally, given the disparate sample sizes between SCTs and datasets, the lack of an observed significant association between a phecode and SCT in our analysis does not definitively indicate that an association does not exist. Rather, the findings we report in this study are the strongest, most consistent phenotypic differences associated with an additional X or Y chromosome, regardless of clinical ascertainment of the SCT.

In conclusion, our PheWAS of 2,769 individuals with SCT from three diverse population-based biobanks— MVP, FinnGen, and UK Biobank—confirmed some previously reported comorbidities enriched across all systems except for neoplasms. Importantly, the observed phenotype is strikingly similar across the three SCTs (47,XXY, 47,XYY, and 47,XXX), independent of the genetic sex and data source, indicating the notable phenotypical implications of having an additional chromosome X or Y in humans.

## Ethics Statement

Patients and control subjects in FinnGen provided informed consent for biobank research, based on the Finnish Biobank Act. Alternatively, separate research cohorts, collected prior the Finnish Biobank Act came into effect (in September 2013) and start of FinnGen (August 2017), were collected based on study-specific consents and later transferred to the Finnish biobanks after approval by Fimea (Finnish Medicines Agency), the National Supervisory Authority for Welfare and Health. Recruitment protocols followed the biobank protocols approved by Fimea. The Coordinating Ethics Committee of the Hospital District of Helsinki and Uusimaa (HUS) statement number for the FinnGen study is Nr HUS/990/2017. The FinnGen study is approved by Finnish Institute for Health and Welfare (permit numbers: THL/2031/6.02.00/2017, THL/1101/5.05.00/2017, THL/341/6.02.00/2018, THL/2222/6.02.00/2018, THL/283/6.02.00/2019, THL/1721/5.05.00/2019 and THL/1524/5.05.00/2020), Digital and population data service agency (permit numbers: VRK43431/2017-3, VRK/6909/2018-3, VRK/4415/2019-3), the Social Insurance Institution (permit numbers: KELA 58/522/2017, KELA 131/522/2018, KELA 70/522/2019, KELA 98/522/2019, KELA 134/522/2019, KELA 138/522/2019, KELA 2/522/2020, KELA 16/522/2020), Findata permit numbers THL/2364/14.02/2020, THL/4055/14.06.00/2020,,THL/3433/14.06.00/2020, THL/4432/14.06/2020, THL/5189/14.06/2020, THL/5894/14.06.00/2020, THL/6619/14.06.00/2020, THL/209/14.06.00/2021, THL/688/14.06.00/2021, THL/1284/14.06.00/2021, THL/1965/14.06.00/2021, THL/5546/14.02.00/2020, THL/2658/14.06.00/2021, THL/4235/14.06.00/202, Statistics Finland (permit numbers: TK-53-1041-17 and TK/143/07.03.00/2020 (earlier TK-53-90-20) TK/1735/07.03.00/2021, TK/3112/07.03.00/2021) and Finnish Registry for Kidney Diseases permission/extract from the meeting minutes on 4th July 2019. The Biobank Access Decisions for FinnGen samples and data utilized in FinnGen Data Freeze 9 include: THL Biobank BB2017_55, BB2017_111, BB2018_19, BB_2018_34, BB_2018_67, BB2018_71, BB2019_7, BB2019_8, BB2019_26, BB2020_1, Finnish Red Cross Blood Service Biobank 7.12.2017, Helsinki Biobank HUS/359/2017, HUS/248/2020, Auria Biobank AB17-5154 and amendment #1 (August 17 2020), AB20-5926 and amendment #1 (April 23 2020) and it’s modification (Sep 22 2021), Biobank Borealis of Northern Finland_2017_1013, Biobank of Eastern Finland 1186/2018 and amendment 22 § /2020, Finnish Clinical Biobank Tampere MH0004 and amendments (21.02.2020 & 06.10.2020), Central Finland Biobank 1-2017, and Terveystalo Biobank STB 2018001 and amendment 25th Aug 2020. The UKBB analyses were conducted using applications 7089, 9905, and 21552.

All methods were carried out in accordance with relevant guidelines and regulations. All experiments were performed in accordance with the UK Biobank Ethics and Governance Framework, as approved by the UK Biobank Ethics and Governance Council, established by the UK Biobank funders, the Wellcome Trust, and the Medical Research Council. The UK Biobank Ethics Advisory Committee continues to monitor the use of the UK Biobank resource, provide guidance on relevant ethical issues, and update ethical policies as necessary. Informed consent was obtained from all UK Biobank participants in accordance with the UK Biobank Ethics and Governance Framework.

All MVP participants provided written informed consent, and the study was approved by the VA Central Institutional Review Board (IRB).

## Acknowledgements

RLH was additionally funded by the VISN-22 VA Center of Excellence for Stress and Mental Health (CESAMH) and National Institute of Aging R01 grants AG050595, ABG074855, AG05064, AG065385. SD was supported by NICHD K23HD092588. DML was supported by the National Institute of Mental Health (K01MH131847). EEL was supported by the National Institutes of General Medical Sciences (NIGMS) 5K12GM093857-11. TBB is supported by Cooperative Studies Program (CSP) #572.

This work was supported using resources and facilities of the Department of Veterans Affairs (VA) Informatics and Computing Infrastructure (VINCI), including data analytics conducted by its Precision Medicine research team and writing support provided by Kathryn Pridgen, which is funded under the research priority to Put VA Data to Work for Veterans (VA ORD 24-D4V-02). This publication does not represent the views of the Department of Veterans Affairs, the National Institutes of Health, or the United States Government.

This research has been conducted using the UK Biobank Resource application number 30782 (PI Peterson). This work was supported by the National Institutes of Health grant R01MH125938 (RE Peterson, AE Gentry, EE Lancaster, M Singh, TB Bigdeli, C Chatzinakos) and the Brain Behavior Research Foundation NARSAD grant 28632 P&S Fund (RE Peterson). The UK Biobank team would like to acknowledge Bradley T. Webb, PhD, of RTI International for his valuable contributions in setting up data access and facilitating data extraction.

The FinnGen project is funded by two grants from Business Finland (HUS 4685/31/2016 and UH 4386/31/2016) and the following industry partners: AbbVie Inc., AstraZeneca UK Ltd, Biogen MA Inc., Bristol Myers Squibb (and Celgene Corporation & Celgene International II Sàrl), Genentech Inc., Merck Sharp & Dohme LCC, Pfizer Inc., GlaxoSmithKline Intellectual Property Development Ltd., Sanofi US Services Inc., Maze Therapeutics Inc., Janssen Biotech Inc, Novartis AG, and Boehringer Ingelheim International GmbH. Following biobanks are acknowledged for delivering biobank samples to FinnGen: Auria Biobank (www.auria.fi/biopankki), THL Biobank (www.thl.fi/biobank), Helsinki Biobank (www.helsinginbiopankki.fi), Biobank Borealis of Northern Finland (https://www.ppshp.fi/Tutkimus-ja-opetus/Biopankki/Pages/Biobank-Borealis-briefly-in-English.aspx), Finnish Clinical Biobank Tampere (www.tays.fi/en-US/Research_and_development/Finnish_Clinical_Biobank_Tampere), Biobank of Eastern Finland (www.ita-suomenbiopankki.fi/en), Central Finland Biobank (www.ksshp.fi/fi-FI/Potilaalle/Biopankki), Finnish Red Cross Blood Service Biobank (www.veripalvelu.fi/verenluovutus/biopankkitoiminta), Terveystalo Biobank (www.terveystalo.com/fi/Yritystietoa/Terveystalo-Biopankki/Biopankki/) and Arctic Biobank (https://www.oulu.fi/en/university/faculties-and-units/faculty-medicine/northern-finland-birth-cohorts-and-arctic-biobank). All Finnish Biobanks are members of BBMRI.fi infrastructure (www.bbmri.fi). Finnish Biobank Cooperative -FINBB (https://finbb.fi/) is the coordinator of BBMRI-ERIC operations in Finland. The Finnish biobank data can be accessed through the Fingenious® services (https://site.fingenious.fi/en/) managed by FINBB.

## Data Availability

The data and code used to generate MVP results are accessible to researchers with MVP data access. Due to VA policy, MVP is currently only accessible to researchers with a funded MVP project (e.g., VA Merit Award, Career Development Award, NIH R01). See https://genhub.va.gov/file/view/897656 for additional information.

For FinnGen, researchers can apply for health data from the Finnish Data Authority Findata (https://findata.fi/en/permits/) and individual-level genotype data available through the Fingenious portal (https://site.fingenious.fi/en/). These resources are hosted by the Finnish Biobank Cooperative FINBB (https://finbb.fi/en/). Access can only be provided for research projects within the scope of the Finnish Biobank Act, which includes health promotion, understanding disease mechanisms or developing medical products or treatment practices. Codes used to perform FinnGen PheWAS and across-biobank meta-analysis are available at https://github.com/dsgelab/Sex-Chromosome-Aneuploidy_PheWAS.

The UK Biobank data utilized in this research is available to, “bona fide researchers for health-related research in the public interest,” through an application process accessible through the UK Biobank website, https://www.ukbiobank.ac.uk/. Codes used to perform UK Biobank PheWAS are available at https://github.com/POPGEM-Lab/UKB-SCT-PheWAS.

### Biobank contact information

MVP.

FinnGen

UK Biobank

