## Supplemental Information for "Prevalence and disease risks for male and female sex chromosome trisomies: a registry-based phenome-wide association study in 1.5 million participants of MVP, FinnGen, and UK Biobank"

**Supplemental Figure 1. Association analysis of disease diagnosis for 345 clinically diagnosed 47,XXY (Klinefelter Syndrome) from a meta-analysis of MVP, FinnGen, and UK Biobank.**

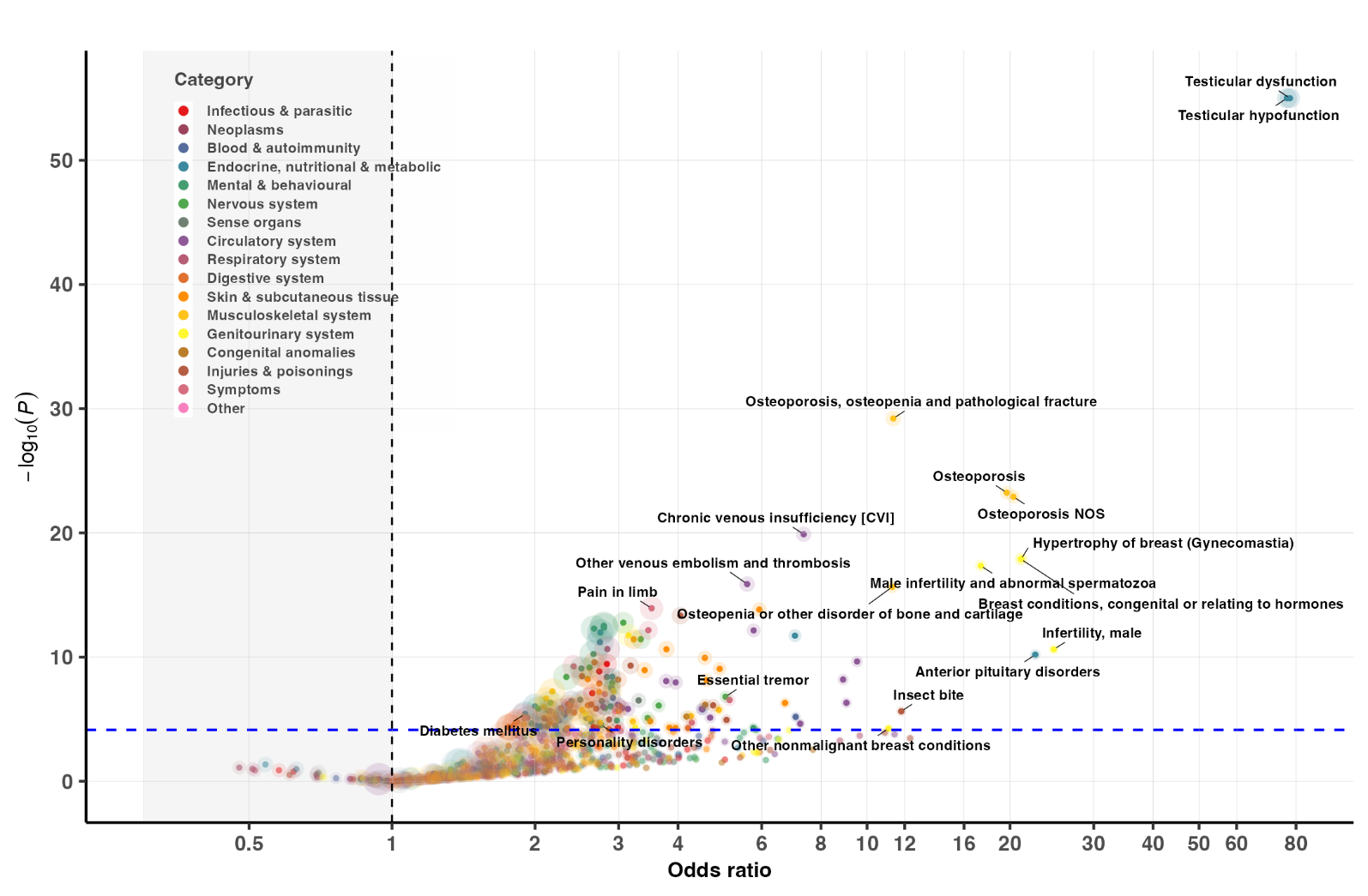

Each 47,XXY case was matched to five 46,XY controls based on birth year and genetic ancestry (when applicable). Logistic regression was used to examine the association between the 47,XXY status and a given phecode within each cohort, followed by a fixed-effect meta-analysis across cohorts. Dashed lines denote the statistically significant threshold after Bonferroni correction (P=0.05/692=7.2e-05); colors represent disease categories; the size of circle reflects the N of disease cases analyzed for each phecode.

**Supplementary Figure 2. Association analysis of disease diagnosis for 974 genetically identified but clinically undiagnosed 47,XXY from a meta-analysis of MVP, FinnGen, and UK Biobank.**

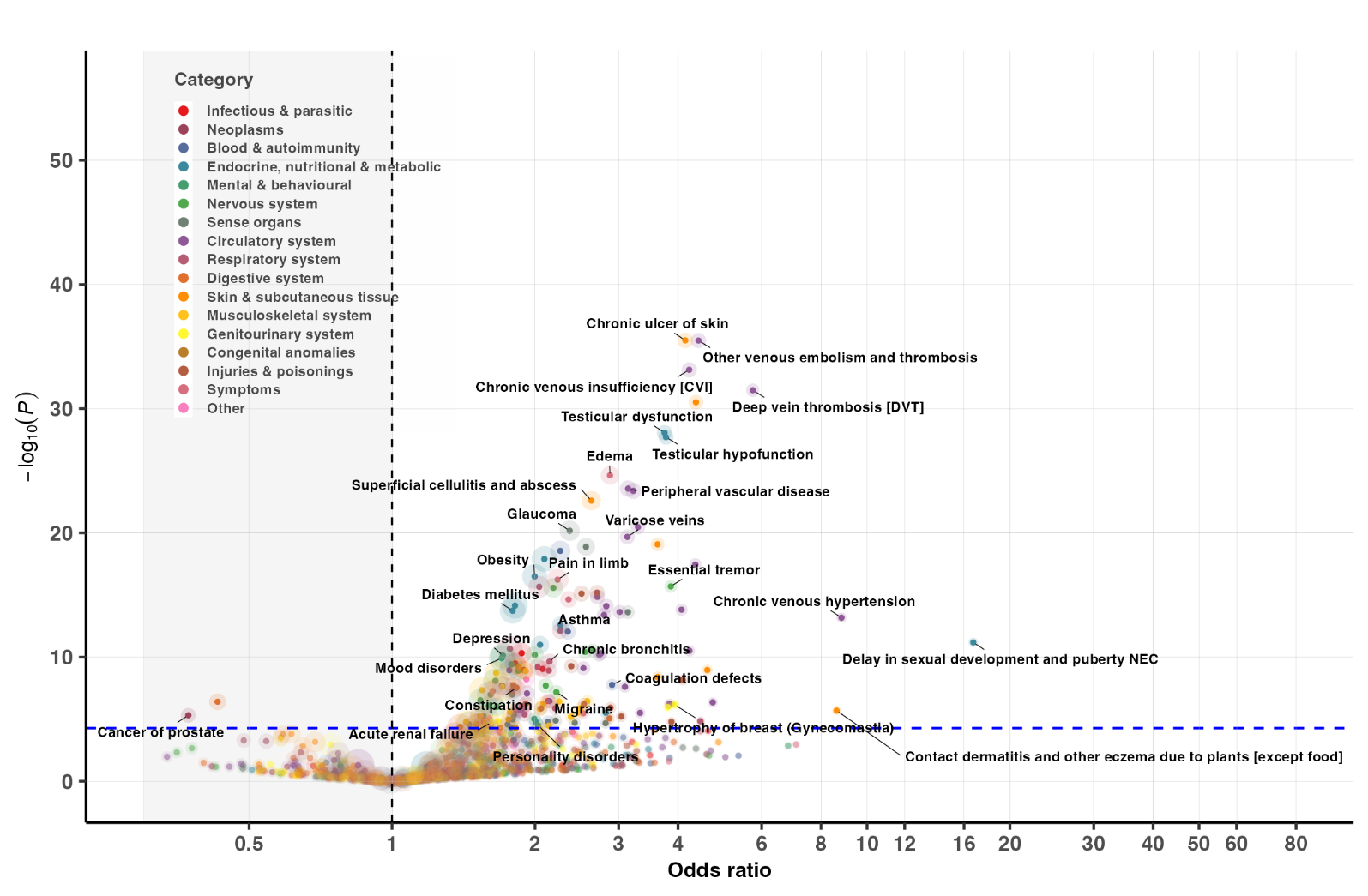

Each 47,XXY case was matched to five 46,XY controls based on birth year and genetic ancestry (when applicable). Logistic regression was used to examine the association between the 47,XXY status and a given phecode within each cohort, followed by a fixed-effect meta-analysis across cohorts. Dashed lines denote the statistically significant threshold after Bonferroni correction (P=0.05/970=5.2e-05); colors represent disease categories; the size of circle reflects the N of disease cases analyzed for each phecode.

| **Supplemental Table 1. Number of analyzable phecodes by karyotype** | | | | | | |
| --- | --- | --- | --- | --- | --- | --- |
|  | **Phecodes in ≥1 dataset** | **Phecodes in ≥2 datasets** | **Significance threshold** | **Significantly different in SCT, N (%)** | **Greater in SCT**  **N (%)** | **Lower in SCT**  **N (%)** |
| **47,XXY** | 1,076 | 495 | 1.0 x 10^-4^ | 183 (37.0%) | 182 (36.8%) | 1 (0.2%) |
| ***Clinically diagnosed*** | 684 | 178 | 2.8 x 10^-4^ | 89 (50.0%) | 89 (50.0%) | 0 |
| ***Clinically undiagnosed*** | 963 | 414 | 1.2 x 10^-4^ | 118 (28.5%) | 117 (28.2%) | 1 (<0.1%) |
| **47,XYY** | 1,007 | 450 | 1.1 x 10^-4^ | 142 (31.6%) | 140 (31.1%) | 2 (0.4%) |
| **47,XXX** | 522 | 222 | 2.3 x 10^-4^ | 62 (27.9%) | 62 (27.9%) | 0 |
| A total of 1,866 phecodes are present in rolled map v1.2. Phecodes present in at least 5 cases and controls were included in this meta-analysis. Significance threshold was determined based on the Bonferroni correction for number of phecodes in ≥2 datasets. | | | | | | |

| **Supplemental Table 2. Proportion of differential phecodes by system category for SCT vs controls** | | | | | | |
| --- | --- | --- | --- | --- | --- | --- |
|  | **XXY** | | **XYY** | | **XXX** | |
|  | **Significant** | **OR>2.0** | **Significant** | **OR>2.0** | **Significant** | **OR>2.0** |
| **All systems** | 182/495 (36.8%) | 85/495 (17.2%) | 140/450 (31.1%) | 96/450 (21.3%) | 62/222 (27.9%) | 94/222 (42.3%) |
| Circulatory System | 29/75 (38.6%) | 21/75 (28.0%) | 30/63 (47.6%) | 23/63 (36.5%) | 12/27 (44.0%) | 19/27 (70.4%) |
| Skin & Subcutaneous | 9/30 (30.0%) | 5/30 (16.7%) | 14/25 (56.0%) | 12/25 (48.0%) | 1/10 (10.0%) | 3/10 (30.0%) |
| Digestive System | 15/67 (22.4%) | 0/67 (0%) | 4/62  (6.5%) | 2/62 (3.2%) | 6/22 (27.2%) | 8/22 (36.4%) |
| Endocrine/Metabolic | 14/30 (46.7%) | 7/30 (23.3%) | 12/28 (42.9%) | 9/28 (32.1%) | 4/15 (26.7%) | 7/15 (46.7%) |
| GU System | 9/33 (27.3%) | 4/33 (12.1%) | 13/34 (38.2%) | 5/34 (14.7%) | 3/28 (10.7%) | 7/28 (25.0%) |
| Hematopoietic | 5/7  (71.4%) | 3/7 (42.9%) | 3/5  (60.0%) | 0/5 (0%) | 1/3  (33.3%) | 1/3 (33.3%) |
| Infectious Diseases | 8/17 (47.1%) | 3/17 (17.6%) | 10/18 (55.6%) | 11/18 (61.1%) | 0/4  (0%) | 3/4 (75.0%) |
| Injuries & Poisonings | 14/33 (42.4%) | 9/33 (27.3%) | 10/33 (30.3%) | 12/33 (36.4%) | 3/10 (30.0%) | 5/10 (50.0%) |
| Mental & Behavioral | 14/33 (42.4%) | 5/33 (15.2%) | 4/27 (14.8%) | 2/27 (7.4%) | 4/14 (28.6%) | 8/14 (57.1%) |
| Musculoskeletal | 16/35 (45.7%) | 6/35 (17.1%) | 4/28 (14.3%) | 0/28 (0%) | 6/24 (25.0%) | 7/24 (29.2%) |
| Neoplasms | 1/19  (5.3%) | 1/19 (5.3%) | 0/20  (0%) | 0/20 (0%) | 0/3  (0%) | 0/3  (0%) |
| Neurologic System | 14/19 (73.7%) | 8/19 (42.1%) | 8/19 (42.1%) | 5/19 (26.3%) | 5/9  (55.6%) | 5/9 (55.6%) |
| Respiratory System | 11/35 (31.4%) | 4/35 (11.4%) | 14/35 (40.0%) | 9/35 (25.7%) | 10/17 (58.9%) | 10 (58.9%) |
| Sense Organs | 11/34 (32.4%) | 4/34 (11.8%) | 8/32 (25.0%) | 4/32 (12.5%) | 1/20  (5.0%) | 5/20 (25.0%) |
| Symptoms | 9/14 (64.3%) | 4/14 (28.6%) | 4/12 (33.3%) | 1/12 (8.3%) | 4/11 (36.4%) | 4/11 (36.4%) |
| A total of 1866 phecodes are present in rolled map v1.2. Phecodes present in at least 5 cases and controls from ≥2 biobanks were included in this meta-analysis. Significance threshold was determined based on the Bonferroni correction for the number of phecodes analyzed. | | | | | | |

**Supplemental Table 3.** See CSV file for meta-analysis results for all three SCTs.

**Supplemental Table 4.** See CSV file for meta-analysis results in XXY stratified by clinical diagnosis status.
